# Effect of Convalescent Plasma on Mortality among Hospitalized Patients with COVID-19: Initial Three-Month Experience

**DOI:** 10.1101/2020.08.12.20169359

**Authors:** Michael J. Joyner, Jonathon W. Senefeld, Stephen A. Klassen, John R. Mills, Patrick W. Johnson, Elitza S. Theel, Chad C. Wiggins, Katelyn A. Bruno, Allan M. Klompas, Elizabeth R. Lesser, Katie L. Kunze, Matthew A. Sexton, Juan C. Diaz Soto, Sarah E. Baker, John R.A. Shepherd, Noud van Helmond, Nigel S. Paneth, DeLisa Fairweather, R. Scott Wright, Rickey E. Carter, Arturo Casadevall, the US EAP COVID-19 Plasma Consortium

**Affiliations:** Department of Anesthesiology and Perioperative Medicine, Mayo Clinic, Rochester, Minnesota; Department of Laboratory Medicine and Pathology, Mayo Clinic, Rochester, Minnesota; Department of Health Sciences Research, Mayo Clinic, Jacksonville, Florida; Department of Cardiovascular Medicine, Mayo Clinic, Jacksonville, Florida; Department of Health Sciences Research, Mayo Clinic, Scottsdale, Arizona; Department of Anesthesiology, Cooper Medical School of Rowan University, Cooper University Health Care, Camden, New Jersey; Department of Epidemiology and Biostatistics, College of Human Medicine, Michigan State University, East Lansing, Michigan; Department of Pediatrics and Human Development, College of Human Medicine, Michigan State University, East Lansing, Michigan; Department of Cardiovascular Medicine, Mayo Clinic, Rochester, Minnesota; Human Research Protection Program, Mayo Clinic, Rochester, Minnesota; Department of Molecular Microbiology and Immunology, Johns Hopkins Bloomberg School of Public Health, Baltimore, Maryland; Department of Internal Medicine, Division of Pulmonary and Critical Care Medicine, Mayo Clinic, Rochester, Minnesota; Department of Internal Medicine, Division of Infectious Diseases, Mayo Clinic, Phoenix, Arizona

## Abstract

**Importance:** Passive antibody transfer is a longstanding treatment strategy for infectious diseases that involve the respiratory system. In this context, human convalescent plasma has been used to treat coronavirus disease 2019 (COVID-19), but the efficacy remains uncertain.

**Objective:** To explore potential signals of efficacy of COVID-19 convalescent plasma.

**Design:** Open-label, Expanded Access Program (EAP) for the treatment of COVID-19 patients with human convalescent plasma.

**Setting:** Multicenter, including 2,807 acute care facilities in the US and territories.

**Participants:** Adult participants enrolled and transfused under the purview of the US Convalescent Plasma EAP program between April 4 and July 4, 2020 who were hospitalized with (or at risk of) severe or life threatening acute COVID-19 respiratory syndrome.

**Intervention:** Transfusion of at least one unit of human COVID-19 convalescent plasma using standard transfusion guidelines at any time during hospitalization. Convalescent plasma was donated by recently-recovered COVID-19 survivors, and the antibody levels in the units collected were unknown at the time of transfusion.

**Main Outcomes and Measures:** Seven and thirty-day mortality.

**Results:** The 35,322 transfused patients had heterogeneous demographic and clinical characteristics. This cohort included a high proportion of critically-ill patients, with 52.3% in the intensive care unit (ICU) and 27.5% receiving mechanical ventilation at the time of plasma transfusion. The seven-day mortality rate was 8.7% [95% CI 8.3%-9.2%] in patients transfused within 3 days of COVID-19 diagnosis but 11.9% [11.4%-12.2%] in patients transfused 4 or more days after diagnosis (p<0.001). Similar findings were observed in 30-day mortality (21.6% vs. 26.7%, p<0.0001). Importantly, a gradient of mortality was seen in relation to IgG antibody levels in the transfused plasma. For patients who received high IgG plasma (>18.45 S/Co), seven-day mortality was 8.9% (6.8%, 11.7%); for recipients of medium IgG plasma (4.62 to 18.45 S/Co) mortality was 11.6% (10.3%, 13.1%); and for recipients of low IgG plasma (<4.62 S/Co) mortality was 13.7% (11.1%, 16.8%) (p=0.048). This unadjusted dose-response relationship with IgG was also observed in thirty-day mortality (p=0.021). The pooled relative risk of mortality among patients transfused with high antibody level plasma units was 0.65 [0.47-0.92] for 7 days and 0.77 [0.63-0.94] for 30 days compared to low antibody level plasma units.

**Conclusions and Relevance:** The relationships between reduced mortality and both earlier time to transfusion and higher antibody levels provide signatures of efficacy for convalescent plasma in the treatment of hospitalized COVID-19 patients. This information may be informative for the treatment of COVID-19 and design of randomized clinical trials involving convalescent plasma.

**Trial Registration:** ClinicalTrials.gov Identifier: NCT04338360

**Key Points:** *Question:* Does transfusion of human convalescent plasma reduce mortality among hospitalized COVID-19 patients?

*Findings:* Transfusion of convalescent plasma with higher antibody levels to hospitalized COVID-19 patients significantly reduced mortality compared to transfusions with low antibody levels. Transfusions within three days of COVID-19 diagnosis yielded greater reductions in mortality.

*Meaning:* Embedded in an Expanded Access Program providing access to COVID-19 convalescent plasma and designed to assess its safety, several signals consistent with efficacy of convalescent plasma in the treatment of hospitalized COVID-19 patients emerged.

## Introduction

Passive antibody transfer, including convalescent plasma or serum, has previously been used to treat infectious diseases that involve the respiratory system ^1-3^. This therapeutic approach was established early in the last century and included widespread use of convalescent plasma for treatment of the 1918 influenza ^4^ In this context, the coronavirus disease 2019 (COVID-19) pandemic has revived interest in the use of convalescent plasma for the treatment of hospitalized patients with COVID-19. Although there is substantial interest in the use of COVID-19 convalescent plasma, the efficacy signals are preliminary ^5,6^.

In response to the global COVID-19 pandemic and need for access to treatments possibly providing benefit while randomized clinical trials were in various stages of development and enrollment, the Mayo Clinic initiated the US Expanded Access Program (EAP) for convalescent plasma, which resulted in widespread use of convalescent plasma to treat COVID-19 in the U.S. The EAP received collaborative and financial support from the Biomedical Advanced Research and Development Authority (BARDA). Although the charter of the EAP was to provide access to and assess the safety of COVID-19 convalescent plasma, we performed exploratory analyses on the efficacy of this agent. We hypothesized, based on historical data that earlier administration of convalescent plasma with high antibody levels would be associated with reduced mortality. To address this hypothesis, we evaluated seven and 30-day mortality in 35,322 hospitalized adults transfused with COVID-19 convalescent plasma by asking two questions. First, was earlier treatment of patients with convalescent plasma after diagnosis of COVID-19 associated with reduced mortality compared to later treatment in the course of disease? Second, were higher antibody levels in the transfused convalescent plasma associated with reduced mortality?

## Methods

### Design and Oversight

As described previously^7,8^, the EAP was a US Government-sponsored, national, pragmatic intervention conducted as a multicenter, open-label protocol in hospitalized adults with COVID-19. All hospitals or acute care facilities in the US and any physician licensed in the US were eligible to participate provided they agreed to adhere to the treatment protocol, FDA, and state regulations.

Mayo Clinic served as the academic research organization conducting the study. The Mayo Clinic Institutional Review Board (IRB) was the central IRB, approved the protocol all modifications, and performed regulatory oversight for all sites and investigators. The principal investigator served as the regulatory sponsor. A Data and Safety Monitoring Board oversaw the safety analyses and advised the regulatory sponsor and the Mayo Clinic IRB on risk. Written informed consent was obtained from the participant or a legally-authorized representative prior to enrollment, except for those patients who necessitated use of an emergency consent process defined in collaboration with the US FDA.

### Participants

Eligible patients were aged 18 years or older, hospitalized with a laboratory confirmed diagnosis of infection with severe acute respiratory syndrome coronavirus 2 (SARS-CoV-2), and had (or were judged by a healthcare provider to be at high risk of progression to) severe or life-threatening COVID-19. Inclusion criteria and the clinical symptoms defining severe or life-threatening COVID-19 are outlined in **Supplement 1**.

### Plasma Collection

Convalescent plasma was obtained from a registered or licensed blood collector, and COVID-19 antibody levels were unknown at the time of plasma collection. Convalescent plasma was donated by COVID-19 survivors with confirmed diagnosis via clinical laboratory test whom were symptom free for 14 days, or more according to standard blood center procedures^9^. An aliquot of plasma or serum was shipped from a subset of blood collection centers for later antibody testing. At the time of collection, each plasma unit was assigned a standardized identifying number (ISBT 128 code) specific to the *donor*, which was used to link antibody levels with study outcomes corresponding to the plasma *recipient(s)*.

### Plasma Transfusion

Compatible COVID-19 convalescent plasma was administered intravenously according to individual institutional protocols. The transfusion dose of COVID-19 convalescent plasma was at least one unit (approximately 200 mL), with the option to administer additional doses if clinically justified.

### Data Entry

Web-based, standardized data reporting surveys were completed to assess the clinical status of patients using the Research Electronic Data Capture system (REDCap, v.9.1.15 Vanderbilt University, Nashville, TN)^10,11^, with FDA authorization, as previously described^7,8^. Given the rapidity at which the EAP was implemented and considering the stress on clinical staff at participating sites during this on-going pandemic, the web-based case reporting forms were designed to optimize convenience. Additionally, although the patient inclusion criteria were specific to hospitalized patients, these criteria were exceptionally broad (**Supplement 1**). While these elements of the EAP may be atypical, they are perhaps understandable in a crisis of the magnitude of the COVID 19 pandemic.

### Antibody testing

Binding antibody levels from sera were tested using the Ortho-Clinical Diagnostics VITROS Anti-SARS-CoV-2 IgG chemiluminescent immunoassay (CLIA) in accordance with manufacturer instructions^12^. The Ortho-Clinical IgG CLIA is a qualitative assay based on a recombinant form of the SARS-CoV-2 spike subunit 1 protein. Results of this assay are based on the sample signal-to-cut-off (S/Co) ratio, with values <1.0 and ≥1.00 corresponding to negative and positive results, The S/Co values reflect relative levels of anti-SARS-CoV-2 antibodies.

### Statistics

The sample size for the EAP was not determined *a priori* and patient accrual has not concluded at the time of this writing. The sample sizes for these analyses varied by the availability of linked antibody data, and in some cases, missing data. For the analyses not associated with antibody data, all transfusions on or before July 4, 2020 were included (i.e., three months after the first confirmed transfusion in the EAP). The database was locked for this study report on August 5, 2020 to allow all included patients to have up to 30 days of follow up after transfusion. For the subset of patients with remnant samples suitable for antibody analysis, all patients matched by the standardized identifying number (ISBT 128 code) were included, with some caveats detailed below.

Based on insights from the pre-antibiotic era that antibody therapy was most effective when given early^2,13^, our cohort was stratified into categories based on the days from COVID-19 diagnosis to plasma transfusion, including: 0, 1-3, 4-10, and 11 or more days and for some graphical presentations and analyses, dichotomized into 0-3 vs. 4 or more days. The timing of death was recorded within the precision of a calendar day, so adjustments were needed to develop survival estimates. For deaths that occurred on the same day of plasma transfusion, a death indicator representing 0.5 days was assigned. Otherwise, the number of days between plasma transfusion and death was calculated for each patient. Transfused patients were assumed to be alive unless death was recorded via web-based reporting survey.

Given that patients may have had more than one unit of plasma from different donors and the days from diagnosis to transfusion were heterogeneous, decision rules were required for analyses of the antibody data. To control for the potential confounding effects of plasma volume and non-uniform antibody levels between multiple plasma units in the analysis, plasma recipients with a single unit, defined as 150 – 250 mL of plasma, were included in the analysis. Finally, plasma from a single donor may have been fractioned into multiple plasma units and transfused to multiple recipients. The analysis did not adjust for the potential clustering that may have occurred in doing so. For the semi-quantitative Ortho-Clinical IgG assay, low, medium and high relative binding antibody levels were established by setting thresholds for low and high based on the ~20^th^ and ~80^th^ percentiles of the distribution for the S/Co ratios, respectively. Accordingly, the thresholds were set at 4.62 S/Co and 18.45 S/Co.

Unadjusted (crude) mortality and adjusted mortality estimates were constructed. For the unadjusted mortality, or case fatality rate, tabulations of the number of mortality events recorded divided by the total number at risk were computed. Score confidence intervals were estimated. For analysis within subgroups, crude mortality was also estimated by grouping the events on key strata variables (e.g., time to transfusion; time epoch of the study)

With the study being non-randomized and containing multiple sources of possible confounding, adjusted estimates of point mortality were also estimated. Two approaches to adjusting for confounding were implemented. First, the general process of generating crude estimates by strata was used to estimate the relative risk by stratum and then a pooled (common) estimate over all strata was estimated using the Mantel-Haenszel estimator. The second approach for adjusted point estimates was developed as an extension of the methods used for estimating adjusted survival, using a baseline Cox regression model fitted to the data. Without loss of generality, we assumed a single variable of direct interest (e.g., days to transfusion) and a set of covariates to be controlled for within the estimate. Using the ‘conditional’ method for estimating adjusted survival curves^14^, an adjusted estimate of the mortality at Day 7, for example, was obtained. To estimate the confidence interval for the adjusted survival curve, the bootstrap method was used. For each of the bootstrap replicates, the original full data set was used to determine the reference distribution for standardization of the mortality estimate. This approach was extended to provide an estimate of the relative risk over one or more variables of interest. The posterior distribution of potential relative risks was constructed by a Cartesian merge of the posterior adjusted survival estimates for each group. The 2.5^th^ and 97.5^th^ percentiles of this distribution were used as the bootstrap confidence interval for the relative risk. No p-values were provided for this method. The adjustment variables used in these analyses were as follows: time epoch (as shown in **Table 1**), gender, race, age at enrollment (as categories), and indicator variables for having already developed one or more severe COVID-19 conditions (as shown in **Table 1**), being on a ventilator, use of hydroxychloroquine, use of remdesivir, and use of steroids prior to transfusion.

**Table 1.** Patient Characteristics Stratified by Time Period of COVID-19 Convalescent Plasma Transfusion.

|  |  |  |  |  |  |
| --- | --- | --- | --- | --- | --- |
|  | Apr 04 - May 01<br>(N=6,990) | May 01 - Jun 04<br>(N=14,846) | Jun 04 - Jul 04<br>(N=13,486) | Total Patients<br>(N=35,322) | P value |
| **Age at Enrollment (years)** |  |  |  |  | **<0.001** |
| 18 to 39 | 539 (7.7%) | 1,337 (9.0%) | 1,596 (11.8%) | 3,472 (9.8%) |  |
| 40 to 59 | 2,424 (34.7%) | 4,938 (33.3%) | 4,806 (35.6%) | 12,168 (34.4%) |  |
| 60 to 69 | 2,007 (28.7%) | 3,791 (25.5%) | 3,170 (23.5%) | 8,968 (25.4%) |  |
| 70 to 79 | 1,358 (19.4%) | 2,879 (19.4%) | 2,467 (18.3%) | 6,704 (19.0%) |  |
| 80 or older | 662 (9.5%) | 1,901 (12.8%) | 1,447 (10.7%) | 4,010 (11.4%) |  |
| **Gender** |  |  |  |  | **<0.001** |
| Female | 2,546 (36.5%) | 5,961 (40.2%) | 5,489 (40.8%) | 13,996 (39.7%) |  |
| Male | 4,416 (63.4%) | 8,838 (59.7%) | 7,961 (59.1%) | 21,215 (60.2%) |  |
| Undisclosed | 6 (0.1%) | 11 (0.1%) | 11 (0.1%) | 28 (0.1%) |  |
| **Weight Status** |  |  |  |  | **<0.001** |
| Underweight | 69 (1.2%) | 286 (1.9%) | 156 (1.2%) | 511 (1.5%) |  |
| Normal Weight | 1,010 (17.4%) | 2,601 (17.6%) | 1,744 (12.9%) | 5,355 (15.7%) |  |
| Overweight | 1,723 (29.7%) | 4,096 (27.8%) | 3,647 (27.1%) | 9,466 (27.8%) |  |
| Obese | 2,997 (51.7%) | 7,761 (52.6%) | 7,926 (58.8%) | 18,684 (54.9%) |  |
| **Race** |  |  |  |  | **<0.001** |
| White | 3,330 (47.6%) | 7,299 (49.2%) | 7,178 (53.2%) | 17,807 (50.4%) |  |
| Asian | 456 (6.5%) | 628 (4.2%) | 390 (2.9%) | 1,474 (4.2%) |  |
| Black or African American | 1,301 (18.6%) | 2,971 (20.0%) | 2,379 (17.6%) | 6,651 (18.8%) |  |
| Other or Unknown | 1,903 (27.2%) | 3,948 (26.6%) | 3,539 (26.2%) | 9,390 (26.6%) |  |
| **Ethnicity** |  |  |  |  | **<0.001** |
| Hispanic/Latino | 2,391 (34.2%) | 5,297 (35.7%) | 5,875 (43.6%) | 13,563 (38.4%) |  |
| Not Hispanic/Latino | 4,599 (65.8%) | 9,549 (64.3%) | 7,611 (56.4%) | 21,759 (61.6%) |  |
| **Clinical Status** |  |  |  |  |  |
| Current severe or life-threatening COVID-19 | 5,584 (79.9%) | 9,761 (65.7%) | 8,157 (60.5%) | 23,502 (66.5%) | **<0.001** |
| Intensive Care Unit (ICU) care prior to infusion | 4,601 (65.8%) | 7,908 (53.3%) | 5,952 (44.1%) | 18,461 (52.3%) | **<0.001** |
| Mechanical Ventilation prior to infusion | 3,217 (49.9%) | 4,143 (27.9%) | 2,213 (16.4%) | 9,573 (27.5%) | **<0.001** |
| **Severe Risk Factors<sup>a</sup>** |  |  |  |  |  |
| Respiratory failure | 4,063 (72.8%) | 6,352 (65.1%) | 4,760 (58.4%) | 15,175 (64.6%) | **<0.001** |
| Dyspnea | 3,543 (63.4%) | 6,976 (71.5%) | 6,476 (79.4%) | 16,995 (72.3%) | **<0.001** |
| Blood oxygen saturation ≤ 93% | 3,507 (62.8%) | 7,063 (72.4%) | 6,394 (78.4%) | 16,964 (72.2%) | **<0.001** |
| Lung infiltrates > 50% within 24 to 48 hours | 2,415 (43.2%) | 4,151 (42.5%) | 3,015 (37.0%) | 9,581 (40.8%) | **<0.001** |
| Respiratory frequency ≥ 30/min | 2,205 (39.5%) | 4,174 (42.8%) | 3,366 (41.3%) | 9,745 (41.5%) | **<0.001** |
| PaO<sub>2</sub>:FiO<sub>2</sub> ratio < 300 | 1,905 (34.1%) | 3,075 (31.5%) | 1,952 (23.9%) | 6,932 (29.5%) | **<0.001** |
| Multiple organ dysfunction or failure | 1,062 (19.0%) | 1,200 (12.3%) | 560 (6.9%) | 2,822 (12.0%) | **<0.001** |
| Septic shock | 844 (15.1%) | 960 (9.8%) | 475 (5.8%) | 2,279 (9.7%) | **<0.001** |
| **Number of Severe Risk Factors** |  |  |  |  | **<0.001** |
| None | 1,407 (20.1%) | 5,085 (34.3%) | 5,331 (39.5%) | 11,823 (33.5%) |  |
| Limited (1 to 4) | 3,895 (55.7%) | 6,992 (47.1%) | 6,190 (45.9%) | 17,077 (48.3%) |  |
| Many (5+) | 1,688 (24.1%) | 2,769 (18.7%) | 1,965 (14.6%) | 6,422 (18.2%) |  |

|  | Apr 04 - May 01<br>(N=6,990) | May 01 - Jun 04<br>(N=14,846) | Jun 04 - Jul 04<br>(N=13,486) | Total Patients<br>(N=35,322) | P value |
| --- | --- | --- | --- | --- | --- |
| Medications during hospital stay |  |  |  |  |  |
| Angiotensin Receptor Blocker | 397 (5.7%) | 839 (5.7%) | 779 (5.8%) | 2,015 (5.7%) | 0.90 |
| Ace Inhibitor | 467 (6.7%) | 1,130 (7.6%) | 1,023 (7.6%) | 2,620 (7.4%) | **0.032** |
| Azithromycin | 3,811 (54.5%) | 5,717 (38.5%) | 5,456 (40.5%) | 14,984 (42.4%) | **<0.001** |
| Remdesivir | 329 (4.7%) | 4,066 (27.4%) | 6,240 (46.3%) | 10,635 (30.1%) | **<0.001** |
| Steroids | 3,736 (53.4%) | 6,137 (41.3%) | 7,735 (57.4%) | 17,608 (49.8%) | **<0.001** |
| Chloroquine | 33 (0.5%) | 22 (0.1%) | 6 (0.0%) | 61 (0.2%) | **<0.001** |
| Hydroxychloroquine | 4,356 (62.3%) | 2,437 (16.4%) | 245 (1.8%) | 7,038 (19.9%) | **<0.001** |
| Time to Transfusion |  |  |  |  | **<0.001** |
| 0 days | 141 (2.0%) | 598 (4.0%) | 625 (4.6%) | 1,364 (3.9%) |  |
| 1 to 3 days | 1,590 (22.7%) | 5,748 (38.7%) | 6,705 (49.7%) | 14,043 (39.8%) |  |
| 4 to 10 days | 2,843 (40.7%) | 6,244 (42.1%) | 5,271 (39.1%) | 14,358 (40.6%) |  |
| 11+ days | 2,416 (34.6%) | 2,256 (15.2%) | 885 (6.6%) | 5,557 (15.7%) |  |

<sup>a</sup>These data include a subset of the sample (n = 23,502), only those patients that currently have severe or life-threatening COVID-19. Data was not available for Gender (n=83), Weight Status (n=1,306) and Mechanical Ventilation prior to infusion (n=544).

Descriptive statistics are presented as frequencies and percentages. Analytic data are presented as point estimates and 95% confidence intervals. P-values less than 0.05 were considered statistically significant and no correction for multiple testing has been applied to reported p-values. All statistical analyses were completed using R version 3.6.2.

## Results

### Patient Characteristics

Between April 4 and July 4, 2020, 47,047 patients were enrolled in the EAP, of whom 36,226 were transfused with COVID-19 convalescent plasma. Of the 1,959 registered sites with at least one patient enrolled, 1,809 sites had transfused at least one patient (92.3%) and 928 sites had transfused at least ten patients (47.4%), **Figure 1**. Data were included for 35,322 transfused patients with 7-day and 30-day follow-up. Key patient characteristics are presented in **Table 1**, stratified into three groups delineating the time period of the study and COVID-19 pandemic. The data set represented a non-probability sample of hospitalized COVID-19 patients with diverse representation of gender, age, weight status, race, and ethnicity. As shown in **Table 1**, the patients transfused early in the study period (before May 01) were more critically-ill (higher rates of mechanical ventilation, ICU admissions and septic shock), had higher concomitant treatment with hydroxychloroquine and azithromycin, and lower concomitant treatment with remdesivir compared with groups transfused later in the study period.

**Figure 1.**
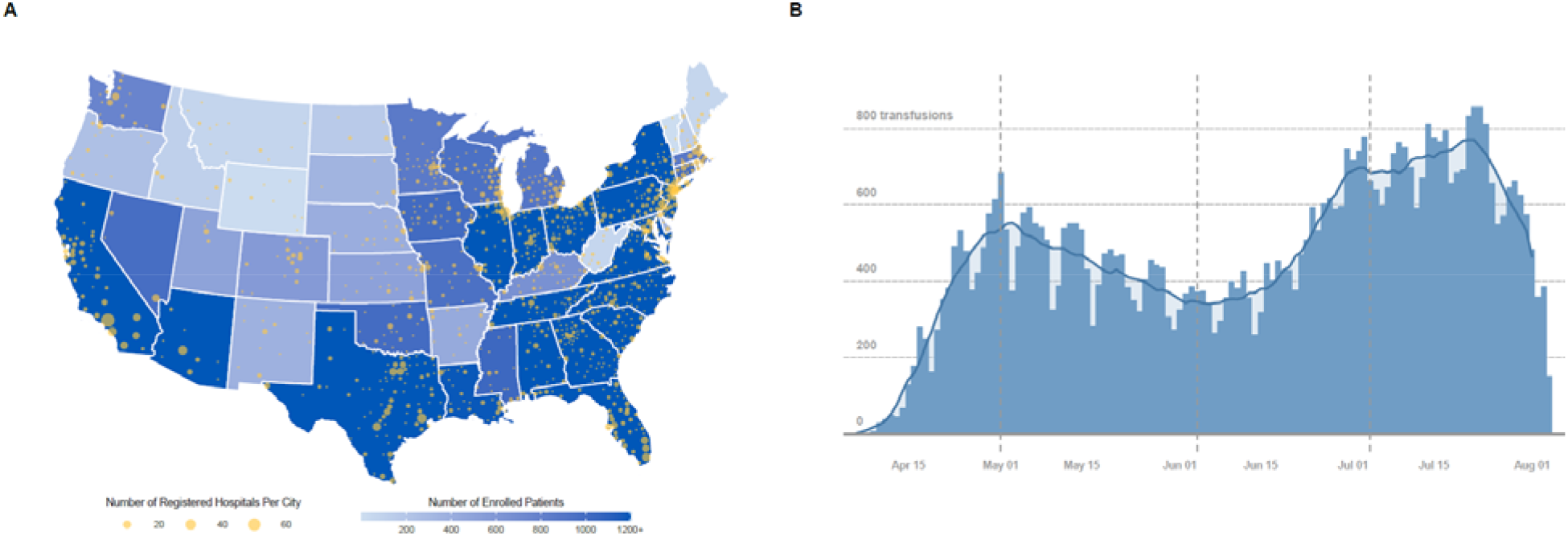
Participation in the US COVID-19 Convalescent Plasma Expanded Access Program (EAP). **A**. Choropleth map displaying the number of cumulatively enrolled patients in the EAP within each state of the contiguous US, with lower enrollment values displayed in a lighter hue and higher enrollment values displayed in a darker hue of blue. Registered acute care facilities are represented as filled yellow circles, with larger circles indicating greater number of registered facilities within the metropolitan area of a city. The choropleth map does not display data from non-contiguous US locations, including registered facilities in Puerto Rico, Hawaii, Alaska, Guam, and Northern Mariana Islands. **B**. The chronological graph represents the number of patients that have received a COVID-19 convalescent plasma transfusion, including daily counts (blue bars) and 7-day average (blue line). The dashed vertical reference lines delineate the three study epochs.

### Unadjusted Analyses

Since the initiation of the EAP, there has been a reduction in both the seven-day crude mortality rate and a pronounced shift of the time to transfusion towards more rapid transfusion of convalescent plasma. The crude seven-day mortality rate was reduced in patients transfused within 3 days (8.7%, 8.3%-9.2%) of COVID-19 diagnosis compared to patients transfused 4 or more days after COVID-19 diagnosis (11.9%, 11.4%-12.3%; *P*<0.001), **Table 2**. Similar trends were seen for unadjusted 30-day mortality. Table 2 presents several additional analyses by risk modifiers (e.g., age and ventilation status at time of transfusion). As a means for controlling for study epoch, the time to transfusion association is presented further stratified by study period. More favorable estimates for mortality were found for all early transfusions (3 or fewer days) across both 7- and 30- day mortality for all three study months (*P*<0.001; **Table 2**).

**Table 2.**
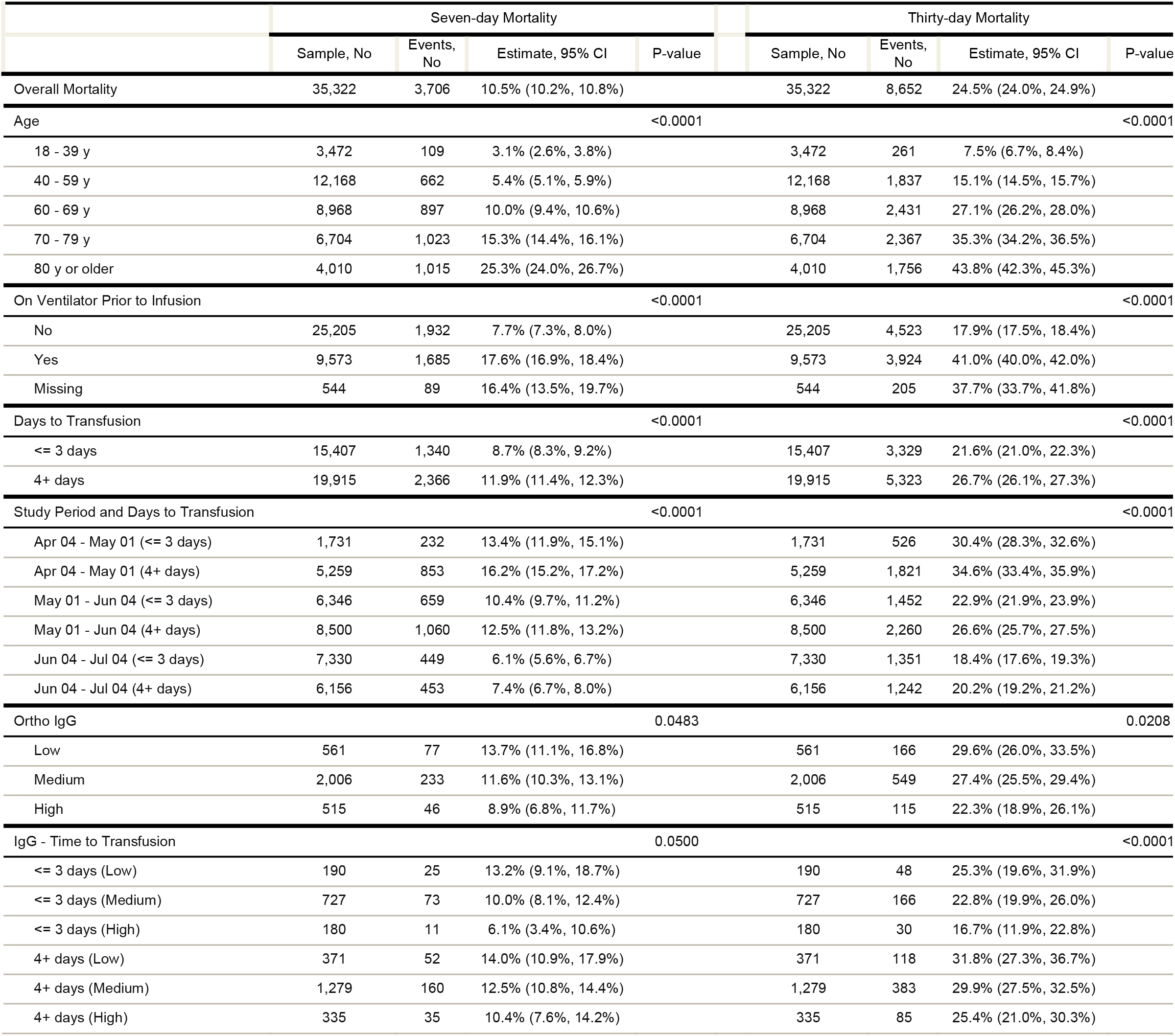
Crude Mortality (7 and 30 day) of patients transfused with CQVID-10 Convalescent Plasma.

|  | Seven-day Mortality |  |  |  | Thirty-day Mortality |  |  |  |
| --- | --- | --- | --- | --- | --- | --- | --- | --- |
|  | Sample, No | Events, No | Estimate, 95% CI | P-value | Sample, No | Events, No | Estimate, 95% CI | P-value |
| Overall Mortality | 35,322 | 3,706 | 10.5% (10.2%, 10.8%) |  | 35,322 | 8,652 | 24.5% (24.0%, 24.9%) |  |
| Age |  |  |  | <0.0001 |  |  |  | <0.0001 |
| 18 - 39 y | 3,472 | 109 | 3.1% (2.6%, 3.8%) |  | 3,472 | 261 | 7.5% (6.7%, 8.4%) |  |
| 40 - 59 y | 12,168 | 662 | 5.4% (5.1%, 5.9%) |  | 12,168 | 1,837 | 15.1% (14.5%, 15.7%) |  |
| 60 - 69 y | 8,968 | 897 | 10.0% (9.4%, 10.6%) |  | 8,968 | 2,431 | 27.1% (26.2%, 28.0%) |  |
| 70 - 79 y | 6,704 | 1,023 | 15.3% (14.4%, 16.1%) |  | 6,704 | 2,367 | 35.3% (34.2%, 36.5%) |  |
| 80 y or older | 4,010 | 1,015 | 25.3% (24.0%, 26.7%) |  | 4,010 | 1,756 | 43.8% (42.3%, 45.3%) |  |
| On Ventilator Prior to Infusion |  |  |  | <0.0001 |  |  |  | <0.0001 |
| No | 25,205 | 1,932 | 7.7% (7.3%, 8.0%) |  | 25,205 | 4,523 | 17.9% (17.5%, 18.4%) |  |
| Yes | 9,573 | 1,685 | 17.6% (16.9%, 18.4%) |  | 9,573 | 3,924 | 41.0% (40.0%, 42.0%) |  |
| Missing | 544 | 89 | 16.4% (13.5%, 19.7%) |  | 544 | 205 | 37.7% (33.7%, 41.8%) |  |
| Days to Transfusion |  |  |  | <0.0001 |  |  |  | <0.0001 |
| <= 3 days | 15,407 | 1,340 | 8.7% (8.3%, 9.2%) |  | 15,407 | 3,329 | 21.6% (21.0%, 22.3%) |  |
| 4+ days | 19,915 | 2,366 | 11.9% (11.4%, 12.3%) |  | 19,915 | 5,323 | 26.7% (26.1%, 27.3%) |  |
| Study Period and Days to Transfusion |  |  |  | <0.0001 |  |  |  | <0.0001 |
| Apr 04 - May 01 (<= 3 days) | 1,731 | 232 | 13.4% (11.9%, 15.1%) |  | 1,731 | 526 | 30.4% (28.3%, 32.6%) |  |
| Apr 04 - May 01 (4+ days) | 5,259 | 853 | 16.2% (15.2%, 17.2%) |  | 5,259 | 1,821 | 34.6% (33.4%, 35.9%) |  |
| May 01 - Jun 04 (<= 3 days) | 6,346 | 659 | 10.4% (9.7%, 11.2%) |  | 6,346 | 1,452 | 22.9% (21.9%, 23.9%) |  |
| May 01 - Jun 04 (4+ days) | 8,500 | 1,060 | 12.5% (11.8%, 13.2%) |  | 8,500 | 2,260 | 26.6% (25.7%, 27.5%) |  |
| Jun 04 - Jul 04 (<= 3 days) | 7,330 | 449 | 6.1% (5.6%, 6.7%) |  | 7,330 | 1,351 | 18.4% (17.6%, 19.3%) |  |
| Jun 04 - Jul 04 (4+ days) | 6,156 | 453 | 7.4% (6.7%, 8.0%) |  | 6,156 | 1,242 | 20.2% (19.2%, 21.2%) |  |
| Ortho IgG |  |  |  | 0.0483 |  |  |  | 0.0208 |
| Low | 561 | 77 | 13.7% (11.1%, 16.8%) |  | 561 | 166 | 29.6% (26.0%, 33.5%) |  |
| Medium | 2,006 | 233 | 11.6% (10.3%, 13.1%) |  | 2,006 | 549 | 27.4% (25.5%, 29.4%) |  |
| High | 515 | 46 | 8.9% (6.8%, 11.7%) |  | 515 | 115 | 22.3% (18.9%, 26.1%) |  |
| IgG - Time to Transfusion |  |  |  | 0.0500 |  |  |  | <0.0001 |
| <= 3 days (Low) | 190 | 25 | 13.2% (9.1%, 18.7%) |  | 190 | 48 | 25.3% (19.6%, 31.9%) |  |
| <= 3 days (Medium) | 727 | 73 | 10.0% (8.1%, 12.4%) |  | 727 | 166 | 22.8% (19.9%, 26.0%) |  |
| <= 3 days (High) | 180 | 11 | 6.1% (3.4%, 10.6%) |  | 180 | 30 | 16.7% (11.9%, 22.8%) |  |
| 4+ days (Low) | 371 | 52 | 14.0% (10.9%, 17.9%) |  | 371 | 118 | 31.8% (27.3%, 36.7%) |  |
| 4+ days (Medium) | 1,279 | 160 | 12.5% (10.8%, 14.4%) |  | 1,279 | 383 | 29.9% (27.5%, 32.5%) |  |
| 4+ days (High) | 335 | 35 | 10.4% (7.6%, 14.2%) |  | 335 | 85 | 25.4% (21.0%, 30.3%) |  |

### Adjusted Analysis including Antibodies

In a subset of 3,082 transfused patients who received only a single unit of plasma (150 – 250 mL), the unadjusted antibody association with mortality is presented in **Table 2**. **Supplemental Table 2** presents the key demographic data by antibody groups (low, medium and high) for these patients. While there were some statistically significant differences among the antibody level groupings, this table shows that patients were well balanced across the antibody level groupings as a whole. The associations of mortality with antibody levels was found at both 7- and 30-days (p<0.05 for both) and when antibody levels were stratified by time to transfusion, a pronounced separation in mortality was found between the extremes of the classification (early transfusion, high antibody levels vs. late transfusion, low antibody levels) albeit the associations for 7-day mortality was at the threshold for statistical significance (p=0.05). **Supplemental Table 2** presents additional estimates of crude mortality on the subset of patients with matched antibody data.

**Figure 2A** presents the adjusted analyses with antibody groupings alone whereas **Figure 2B** presents these same data allowing for the timing of the transfusion to be integrated directly into the analysis. These data demonstrate a clear “dose” dependent relationship of reduced 7-day mortality with the higher antibody levels. **Figure 2C** and **2D** replicate these findings using 30-day mortality data. While some confidence intervals include the null value of relative risk of 1.0, the magnitude of relative risks, particularly after adjustment, is an important finding of the study.

**Figure 2.**
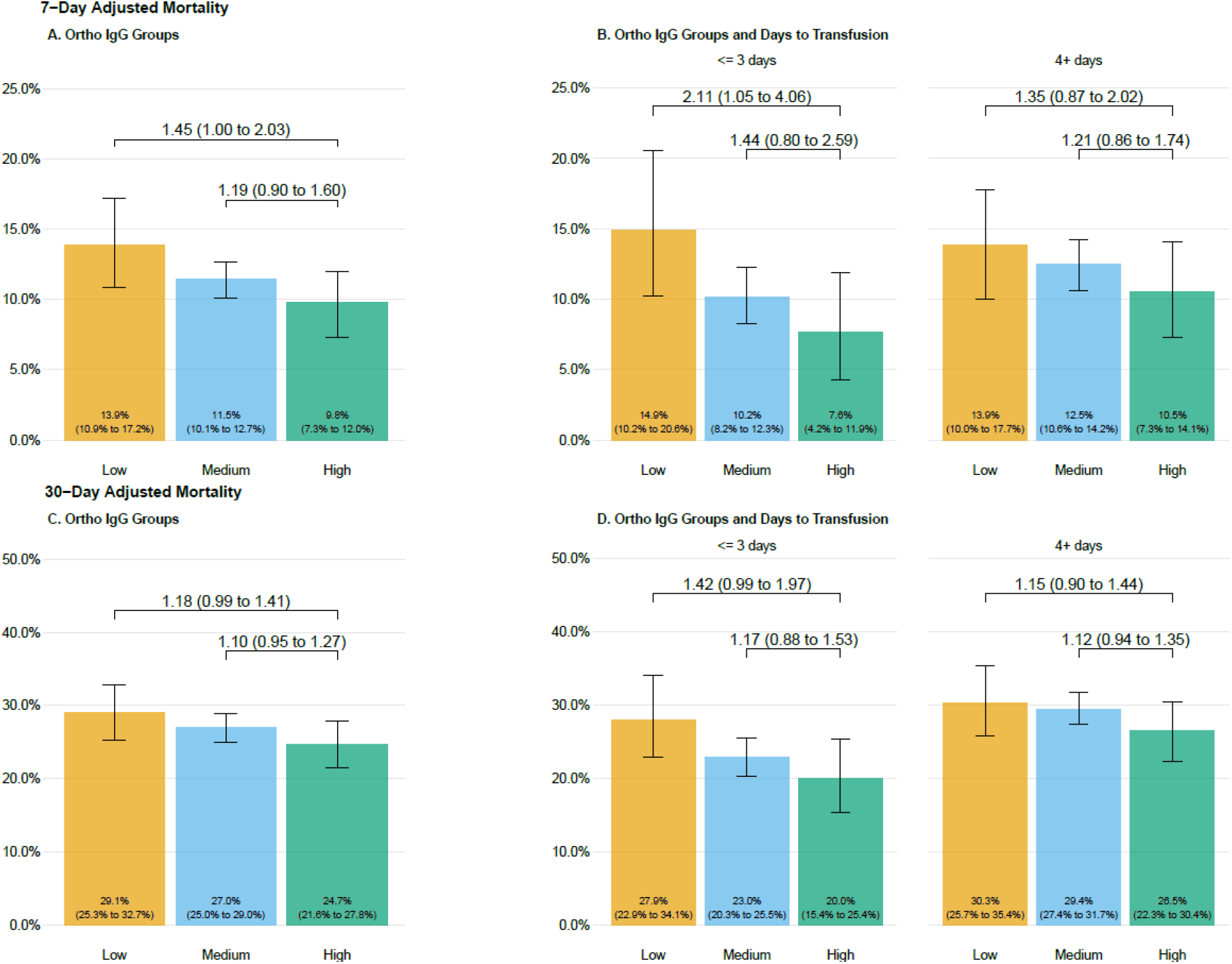
Seven day (A, B) and 30-day (C, D) adjusted mortality stratified by antibody groupings in patients transfused with COVID-19 convalescent plasma. Adjusted mortality rate is presented on the vertical axis, and the height of each bar graph represents adjusted mortality with 95% confidence interval denoted. Data are stratified by groupings of antibody levels with semiquantitative groupings of low (<4.62 S/Co, orange bars), medium (4.62 to 18.45 S/Co, blue bars) and high (> 18.45 S/Co, green bars). Values presented as text within the boxes are the estimated adjusted mortality rates. Values connecting various categories shown with the overbraces are bootstrapped estimates of relative risk and 95% bootstrap confidence intervals. Refer to the methods for the variables in the adjustment and the calculation of the relative risks.

**Figure 3** presents an alternate analytical approach to estimate the effect of the antibody levels. The stratified Mantel-Haenszel approach estimates the relative risk for both 7- and 30-day mortality for patient profiles in the analysis. This stratification approach provides direct analytical control for the potential confounders as each row in the figure represents homogeneity with respect to the factors listed. Overall, there is a consistent signal of a protective effect of the high antibody levels across the strata. The pooled, or common, relative risk for 7-day and 30-day mortality were 0.65 (0.47 to 0.92) and 0.77 (0.63 to 0.94). For this analysis only patients transfused with units containing antibody levels over 18.45 S/Co or less than 4.62 S/Co were included.

**Figure 3.**
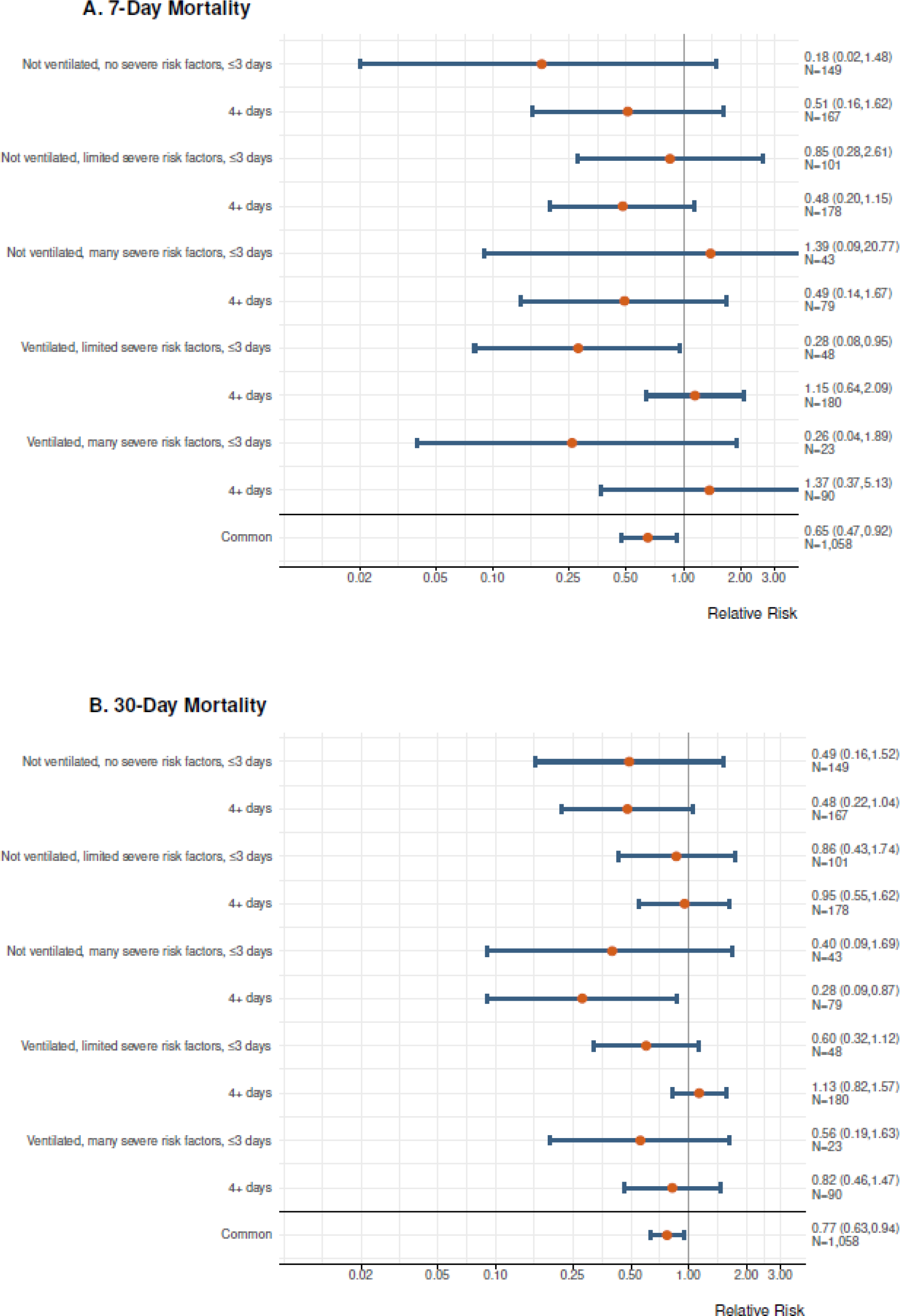
Forest plots of relative risks for 7-day (A) and 30-day (B) mortality for high versus low antibody concentration. Each row in the figure represents 10 mutually exclusive categorizations of patients transfused with convalescent plasma with measured antibody levels. Estimates are the relative risk for mortality for patients who received convalescent plasma with IgG S/co > 18.45 vs. patients that received < 4.62 S/Co. Patients that received units with IgG S/Co values between 4.62 and 18.45 are not included in this analysis as the planned comparison was to highlight the potential efficacy of high IgG containing units vs. units with low levels of detectable antibodies. The bottom row in each figure represents the common (pooled) estimate based on the Mantel-Haenszel estimator. The number of severe risk factors was categorized as none (n=0), limited (n=1 – 4) or many (5 or more), as defined in **Table 1**.

## Discussion

In our cohort of over 35,000 hospitalized patients with COVID-19, several signals consistent with effectiveness for convalescent plasma were observed in a broad sample of acute care facilities across the US. Earlier use of convalescent plasma was associated with lower observed rates of 7-day and 30-day mortality. The use of convalescent plasma with higher antibody levels was associated with reduced 7-day and 30-day mortality. These findings were supported by two different analytical methods used to control for confounding. The finding of a dose response between antibody levels and reduction in mortality provides strong evidence that specific antibody is the active agent in convalescent plasma for treatment of COVID-19. All data considered as a whole, these findings are consistent with the notion that the quality and manner in which convalescent plasma is administered to patients hospitalized with COVID-19 may reduce mortality.

Given the historical efficacy of passive antibody therapy for infectious diseases, the primary objective of the EAP was to facilitate access to convalescent plasma for hospitalized COVID-19 patients across the US. The other major goal was to assess safety. With these goals met^7,8^, we analyzed the data from 1,809 sites and noted there was variability in time to transfusion after diagnosis. Initially, we had no information about the antibody levels in the convalescent plasma being administered. These factors provided elements of inherent randomization in the data collected and formed the basis of an exploratory analysis for signals associated to efficacy. They are also consistent with key principles of antibody therapy recognized during the heyday of this treatment modality in the 1920s and 30s^15,16^, supporting their use as a framework to explore the efficacy of convalescent plasma in COVID-19.

### Time to Treatment

Both 7-day and 30-day mortality adjusted for disease severity and demographic factors were reduced in patients transfused within 3 days of COVID-19 diagnosis compared to patients transfused 4 or more days after COVID-19 diagnosis. Additionally, the declining week-to-week trends in crude mortality (as previously observed^7^) were temporally associated with more rapid treatment. Prior to the antibiotic era, treatment of respiratory infections with antibody therapy was most effective if initiated within three days of hospitalization. Thus, we used a similar timeframe, relative to date of diagnosis rather than hospitalization, for stratifying the current data. Along similar lines, 7 and 30 day survivors received on average higher volumes of plasma in unadjusted analyses. This is of interest because we had no knowledge of the volume of plasma which might constitute an efficacious dose prior to beginning this study.

### Antibody Assessment

Seven and 30 day mortality rates were reduced in patients who received plasma with higher antibody levels. This finding is more limited than the time data as only a subset of the plasma units had remnant samples preserved that were suitable for assaying antibody levels. The survival benefit became more pronounced when the analysis was restricted to less severely ill patients treated early. Because of the multifactorial nature of antibody-mediated effects and the potential for other disease modifying factors to be present in convalescent plasma, further assay development to more fully characterize the mechanisms in which plasma confers anti-viral properties is warranted. We also note that there was no evidence of worsening outcomes or increased mortality in patients treated with very high antibody levels indicating that antibody dependent enhancement of disease was unlikely. Finally, it is important to recognize that the antibody levels we obtained were on repurposed remnant biospecimens collected for blood banking quality assurance. Thus there was potential variability in a number of factors related to biospecimen handling and storage that might influence the measurement of antibody levels in the specimens available to us. Of note adjusted 30- day mortality was 30% in patients treated with plasma with low antibody levels (IgG) 4 or more days after COVID-19 diagnosis. By contrast 30-day mortality was 20% in patients treated within 3 days of diagnosis with plasma with high antibody levels. The pooled estimate from the stratified analysis estimated a 23% relative reduction in mortality at 30 days across a wide range of sub-strata within the study. This reduction in mortality is similar to that observed in a number of small randomized trials and retrospective matched control studies^17^.

### Limitations

The design of the EAP has been criticized because it was not a randomized placebo, controlled trial (RCT)^18^. We started the EAP in late March 2020. It was designed to provide access to convalescent plasma largely at hospitals and acute care facilities that were not already part of a RCT or did not have the infrastructure to support complex RCTs. We also envisioned modest total enrollment and our original IRB approval was for 5,000 patients. In this context, our primary goal was to report on the safety of convalescent plasma and to perform an exploratory analysis for potential signals of efficacy. As described earlier, the EAP was a pragmatic study design, organized to allow routine clinical care to dictate the timing and administration of plasma with the collection of real world data. We did not prespecify which medications patients should be on to participate. The enrollment and data collection forms were streamlined to make participation easy for sites engulfed in the work of a pandemic. The use of a central, academic IRB allowed for consistent data evaluation and oversight. We streamlined PI credentialing and IRB reliance processes. All forms were web-based at a time when some believed that SARS-CoV-2 might be transmitted via paper contaminated with the virus. We did not randomly assign treatment strategies or use of adjunctive medications. Nonetheless, there were some elements of randomization or pseudo-randomization in our study. Physicians could choose the timing of convalescent plasma, the number of units administered, any repeat therapies and whether ICU or mechanically ventilated patients were included. Furthermore, the degree of immune activity within the units of convalescent plasma (i.e. specific IgG levels) was not known. It was assumed that patients would receive plasma with low, medium and high antibody levels in a pseudo-randomized manner and that would enable assessment of efficacy.

We acknowledge that RCTs produce evidence of the highest quality in most but not all clinical situations. RCTs can occur when a number of specific criteria are present which allow their conduct. First, RCTs necessitate a stable supply of investigational product (i.e. convalescent plasma) or placebo/comparator which can be pre-positioned at all participating sites. The supply of convalescent plasma in April was not sufficient for such collection and pre-positioning. Second, RCTs require sufficient numbers of sites which have an appropriate patient base to approach for the study. The COVID-19 pandemic has migrated across different US regions every few weeks, making it challenging to predict where sites should be selected and prepared for a RCT. Third, sites must be validated and activated. This work requires training of the investigators and study team members as well as typically on-site visits. The crises of the COVID-19 pandemic were not compatible with these site training and activation activities; travel within the US has been restricted and staff sent to activate sites would likely have been quarantined for two weeks before being able to go to another region to activate sites. Fourth, the very nature of a RCT requires subject willingness to be randomized to active treatment or placebo or a comparator agent. There was no consensus in April nor is there a global consensus now regarding what would be an appropriate placebo-control to use. Fifth, many COVID-19 patients would likely have been distrustful of being randomized to a placebo based upon historical precedent. Sixth, the number of sites who could have participated in a RCT is limited; who was the appropriate ethical entity to pick those sites and to exclude other sites? Our design allowed any willing hospital, PI and patient to be included in the pragmatic, real-world data study. Finally, there were ongoing small RCTs when we started this program. Physicians, hospitals and patients have the choices of this program versus a RCT. It is clear that over 90,000 patients and over 10,000 physicians elected to participate in the pragmatic, real-world evidence study design. We did not indicate our study would prove efficacy or even offer potential help. It was clear that it was a research investigation and informed consent was obtained in all subjects prior to the transfusion of plasma. Perhaps the current design can inform trialists and RCT advocates of the importance of study designs which are easy and simple to join/enroll and which make the workload of participation as easy and clinically relevant as possible.

### Conclusion

The relationships between mortality and both time to plasma transfusion, and antibody levels provide a signature that is consistent with efficacy for the use of convalescent plasma in the treatment of hospitalized COVID-19 patients.

### Disclaimer

The views and opinions expressed in this manuscript are those of the authors and do not reflect the official policy or position of the US Department of Health and Human services and its agencies including the Biomedical Research and Development Authority and the Food and Drug Administration, as well as any agency of the U.S. government. Assumptions made within and interpretations from the analysis are not reflective of the position of any US government entity.

## Data Availability

Data not available.

## Acknowledgements

We thank the dedicated members of the US Convalescent Plasma Expanded Access Program team— Machiko Anderson, Supriya Behl, Lori Bergstrom, Zachary Buchholtz, Brian Butterfield, Isha Chekuri, Joshua Culberson, Grant Dubbels, Adam Eggert, Ree Erickson, Rebekah Frost, Daniel Gaz, Winston Guo, Starr Guzman, Karina Hex, Vidhu Joshi, Megan Knudson, Tessa Kroeninger, Frances Lynch, Tim Miksch, Lisa Muenkel, Ryan Oldenburg, Amy Olofson, Laura Pacheco-Spann, Dr. Kelly Paulson, Dr. Sumedha Penheiter, Melanie Peterson, Katrina Pierce, Michaela Pletsch, Nicloas Saikali, Jeffrey Schmoll, Pamela Skaran, Lindsay Stromback, Edward Swaray, Morgan Swope, Kristine Tree, Joe Wick, Janelle Worthington. We thank the members of the Mayo Clinic Institutional Review Board, the Mayo Clinic Office of Human Research Protection, the Mayo Clinic Office of Research Regulatory support and in particular Mark Wentworth, the Executive Dean of Research at Mayo Clinic Dr. Gregory Gores and the CEO of Mayo Clinic Dr. Gianrico Farrugia for their support and assistance, and the independent Data and Safety Monitoring Board for their work and oversight of the Expanded Access Program— Dr. Allan S. Jaffe (chair), Dr. David O. Warner, Dr. William G. Morice II, Dr. Paula J. Santrach, Dr. Robert L. Frye, Dr. Lawrence J Appel, Dr. Taimur Sher. We thank the members of the National COVID-19 Convalescent Plasma Project (http://ccpp19.org) for their intellectual contributions and support. We thank the participating medical centers and medical teams, and blood centers for their rigorous efforts necessary to make this program possible. We also thank the donors for providing COVID-19 convalescent plasma.

## Contract and Grant Support

This project has been funded in part with Federal funds from the Department of Health and Human Services; Office of the Assistant Secretary for Preparedness and Response; Biomedical Advanced Research and Development Authority under Contract No. 75A50120C00096. Additionally, this study was supported in part by National Center for Advancing Translational Sciences (NCATS) grant UL1TR002377, National Heart, Lung, and Blood Institute (NHLBI) grant 5R35HL139854 (to MJJ), National Institute of Diabetes and Digestive and Kidney Diseases (NIDDK) 5T32DK07352 (to JWS and CCW), Natural Sciences and Engineering Research Council of Canada (NSERC) PDF-532926-2019 (to SAK), National Institute of Allergy and Infectious Disease (NIAID) grants R21 AI145356, R21 AI152318 and R21 AI154927 (to DF), R01 AI152078 9 (to AC), National Heart Lung and Blood Institute RO1 HL059842 (to AC), National Institute on Aging (NIA) U54AG044170 (to SEB), Schwab Charitable Fund (Eric E Schmidt, Wendy Schmidt donors), United Health Group, National Basketball Association (NBA), Millennium Pharmaceuticals, Octapharma USA, Inc, and the Mayo Clinic.

## Supplement 1

### Trial Protocol

#### 1 | Study Objectives

Convalescent plasma is a potential disease altering therapy for hospitalized patients with COVID-19 infections. There is strong historical precedence for its use in respiratory infections suggesting it may be effective in the treatment of COVID-19. Additionally, the administration of convalescent plasma is considered well-tolerated and safe, both historically and within the context of the current COVID-19 pandemic.

##### 1.2 | Primary Objectives

The primary outcome of this Expanded Access Program was to provide access to COVID-19 convalescent plasma, assessed as the availability of convalescent plasma.

##### 1.3 | Secondary Objectives

The secondary outcome of this Expanded Access Program was to determine the safety of transfusion of COVID-19 convalescent plasma assessed as the case-rate and relatedness of serious adverse events.

##### 1.4 | Tertiary Objectives

The tertiary outcome of this Expanded Access Program was to explore the efficacy of transfusion of COVID-19 convalescent plasma.

#### 2 | Study Intervention

This Expanded Access Program was a national, pragmatic intervention conducted as a multicenter, open-label protocol in hospitalized adults with COVID-19. All patients received the study intervention (COVID-19 convalescent plasma transfusion). Primary study endpoints included:

1. Hospital discharge
2. Death
3. 30 days of observation after COVID-19 convalescent plasma transfusion

##### 2.1 | Study Intervention Description

Compatible COVID-19 convalescent plasma was administered intravenously according to accepted transfusion guidelines used for fresh frozen plasma.

##### 2.2 | Dosing and Administration

For practical purposes in the current outbreak, one – two units of compatible COVID-19 convalescent plasma were administered. Convalescent plasma was obtained from a registered or licensed blood collector and was collected preferably by apheresis or, if necessary, by conventional methods. Individual institutional guidelines for the administration of plasma were followed, including the use of any premedications, such as acetaminophen or diphenhydramine.

##### 2.3 | Preparation and Packaging

Compatible convalescent plasma units were obtained from a registered or licensed blood collector following registration of a patient under the auspices of the Expanded Access Program. COVID-19 convalescent plasma was supplied as an investigational blood product for the treatment of COVID-19. The plasma container label of the COVID-19 convalescent plasma unit included the following statement, “Caution: New Drug – Limited by Federal (or United States) law to investigational use.” (21 CFR 312.6(a)).

#### 3 | Research Population

Eligible patients for this Expanded Access Program were identified by their treating providers. The patient inclusion criteria were specific to hospitalized patients, these criteria were exceptionally broad.

##### 3.1 | Inclusion Criteria

**Supplemental Table 1.**
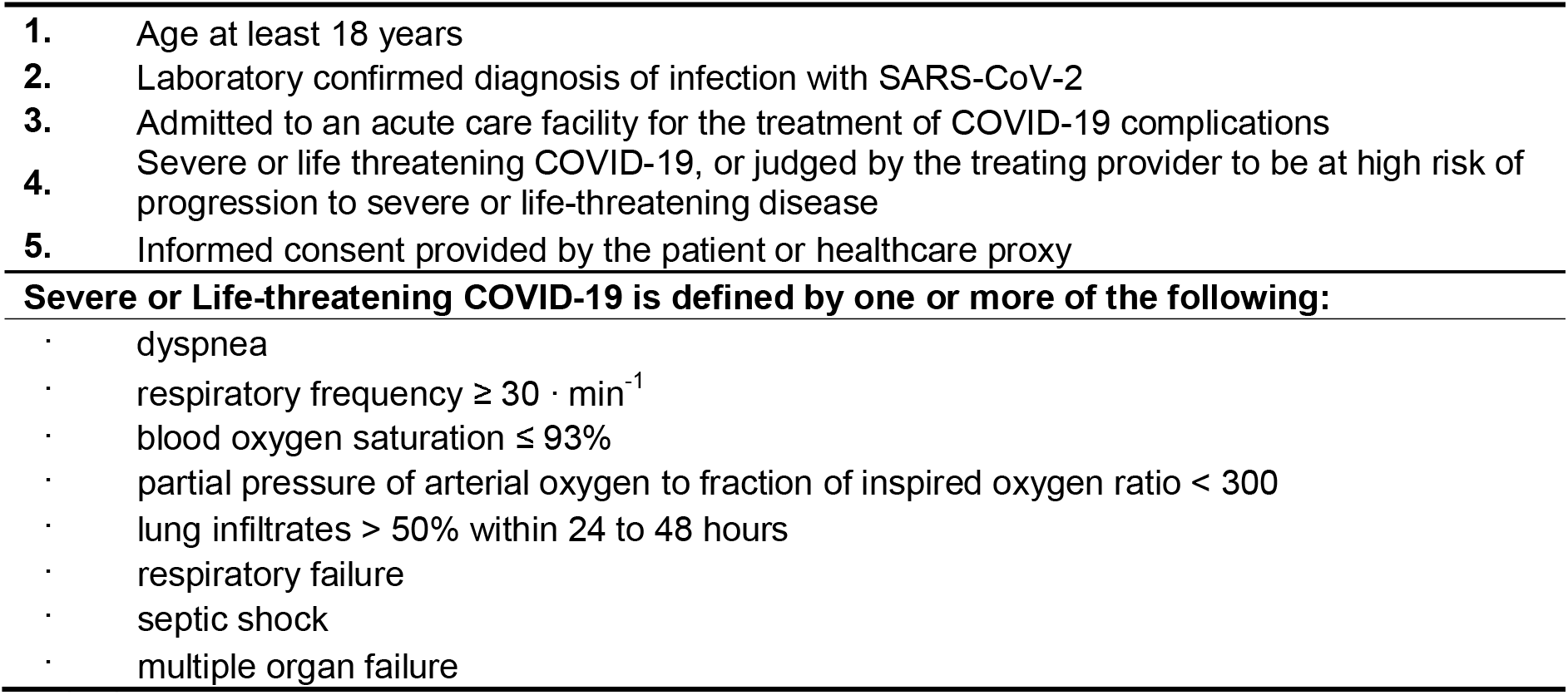
Inclusion Criteria

##### 3.2 | Exclusion Criteria

None.

## Supplement 2

**Supplemental Table 2.**
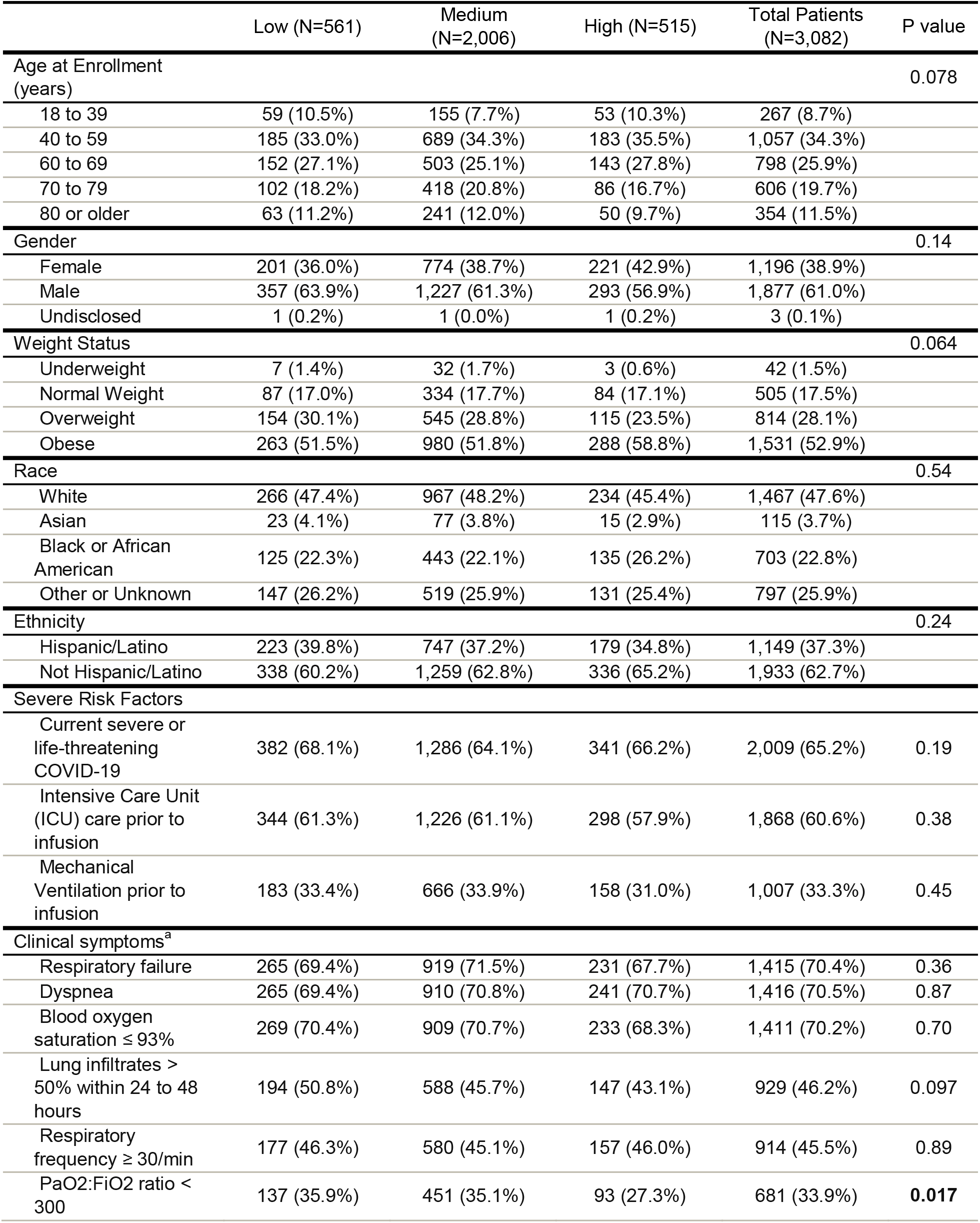
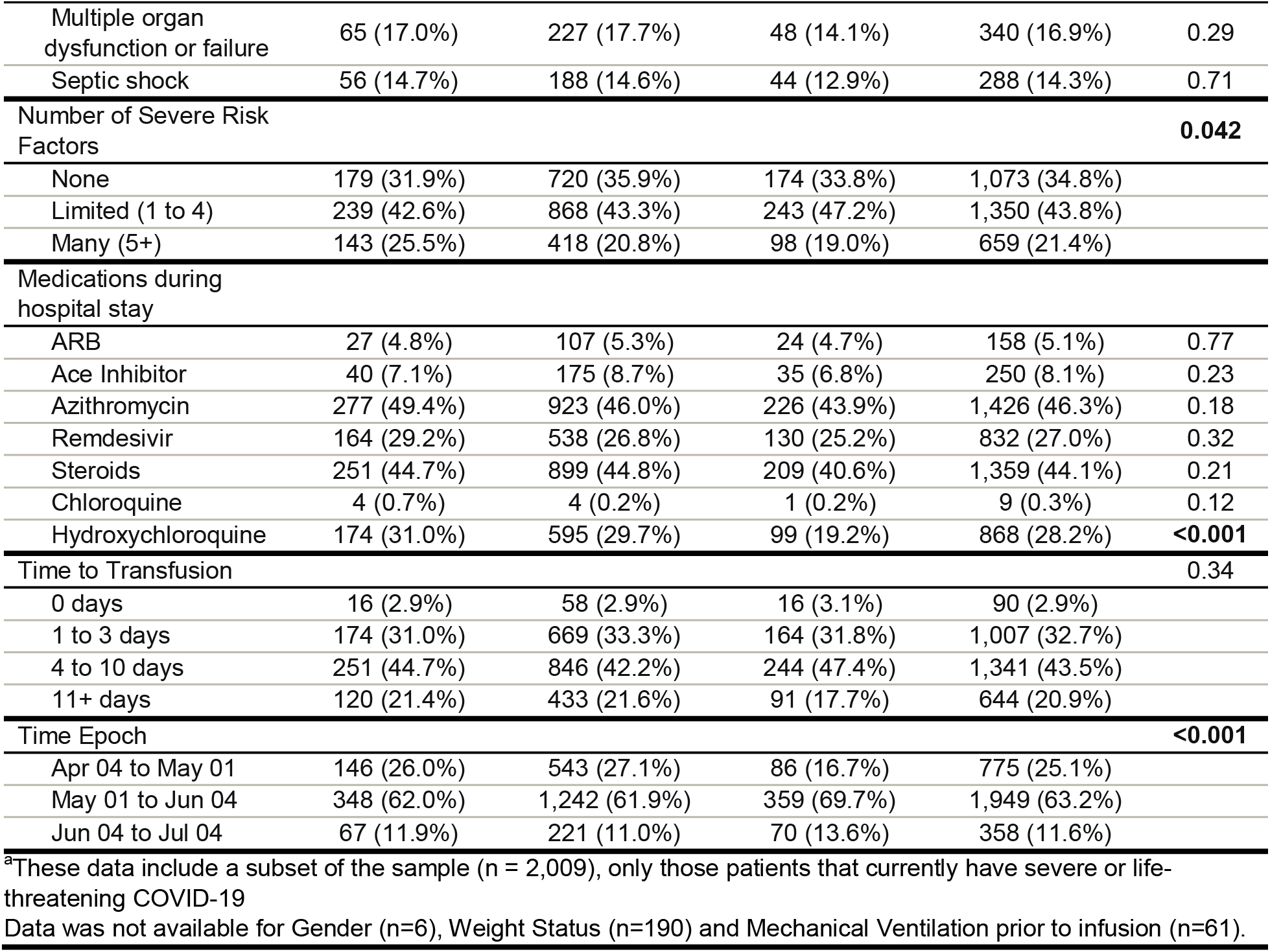
Patient Characteristics Stratified by IgG.

**Supplemental Table 3.** Crude Mortality (7 and 30 day) of patients with IgG transfused with C0VID-10 Convalescent Plasma.

|  | Seven-day Mortality |  |  |  | Thirty-day Mortality |  |  |  |
| --- | --- | --- | --- | --- | --- | --- | --- | --- |
|  | Sample, No | Events, No | Estimate, 95% CI | P-value | Sample, No | Events, No | Estimate, 95% CI | P-value |
| Overall Mortality | 3,082 | 356 | 11.6% (10.5%, 12.7%) |  | 3,082 | 830 | 26.9% (25.4%, 28.5%) |  |
| Age |  |  |  | <0.0001 |  |  |  | <0.0001 |
| 18 - 39 y | 267 | 10 | 3.7% (2.0%, 6.8%) |  | 267 | 27 | 10.1% (7.0%, 14.3%) |  |
| 40 - 59 y | 1,057 | 83 | 7.9% (6.4%, 9.6%) |  | 1,057 | 187 | 17.7% (15.5%, 20.1%) |  |
| 60 - 69 y | 798 | 89 | 11.2% (9.2%, 13.5%) |  | 798 | 243 | 30.5% (27.4%, 33.7%) |  |
| 70 - 79 y | 606 | 97 | 16.0% (13.3%, 19.1%) |  | 606 | 217 | 35.8% (32.1%, 39.7%) |  |
| 80 y or older | 354 | 77 | 21.8% (17.8%, 26.3%) |  | 354 | 156 | 44.1% (39.0%, 49.3%) |  |
| On Ventilator Prior to Infusion |  |  |  | <0.0001 |  |  |  | <0.0001 |
| No | 2,014 | 170 | 8.4% (7.3%, 9.7%) |  | 2,014 | 382 | 19.0% (17.3%, 20.7%) |  |
| Yes | 1,007 | 177 | 17.6% (15.4%, 20.0%) |  | 1,007 | 421 | 41.8% (38.8%, 44.9%) |  |
| Missing | 61 | 9 | 14.8% (8.0%, 25.7%) |  | 61 | 27 | 44.3% (32.5%, 56.7%) |  |
| Days to Transfusion |  |  |  | 0.0371 |  |  |  | <0.0001 |
| <= 3 days | 1,097 | 109 | 9.9% (8.3%, 11.8%) |  | 1,097 | 244 | 22.2% (19.9%, 24.8%) |  |
| 4+ days | 1,985 | 247 | 12.4% (11.1%, 14.0%) |  | 1,985 | 586 | 29.5% (27.6%, 31.6%) |  |
| Study Period and Days to Transfusion |  |  |  | 0.0470 |  |  |  | <0.0001 |
| Apr 04 - May 01 (<= 3 days) | 138 | 14 | 10.1% (6.1%, 16.3%) |  | 138 | 36 | 26.1% (19.5%, 34.0%) |  |
| Apr 04 - May 01 (4+ days) | 637 | 95 | 14.9% (12.4%, 17.9%) |  | 637 | 219 | 34.4% (30.8%, 38.2%) |  |
| May 01 - Jun 04 (<= 3 days) | 773 | 77 | 10.0% (8.0%, 12.3%) |  | 773 | 172 | 22.3% (19.5%, 25.3%) |  |
| May 01 - Jun 04 (4+ days) | 1,176 | 137 | 11.6% (9.9%, 13.6%) |  | 1,176 | 327 | 27.8% (25.3%, 30.4%) |  |
| Jun 04 - Jul 04 (<= 3 days) | 186 | 18 | 9.7% (6.2%, 14.8%) |  | 186 | 36 | 19.4% (14.3%, 25.6%) |  |
| Jun 04 - Jul 04 (4+ days) | 172 | 15 | 8.7% (5.4%, 13.9%) |  | 172 | 40 | 23.3% (17.6%, 30.1%) |  |
| Ortho IgG |  |  |  | 0.0483 |  |  |  | 0.0208 |
| Low | 561 | 77 | 13.7% (11.1%, 16.8%) |  | 561 | 166 | 29.6% (26.0%, 33.5%) |  |
| Medium | 2,006 | 233 | 11.6% (10.3%, 13.1%) |  | 2,006 | 549 | 27.4% (25.5%, 29.4%) |  |
| High | 515 | 46 | 8.9% (6.8%, 11.7%) |  | 515 | 115 | 22.3% (18.9%, 26.1%) |  |
| IgG - Time to Transfusion |  |  |  | 0.0500 |  |  |  | <0.0001 |
| <= 3 days (Low) | 190 | 25 | 13.2% (9.1%, 18.7%) |  | 190 | 48 | 25.3% (19.6%, 31.9%) |  |
| <= 3 days (Medium) | 727 | 73 | 10.0% (8.1%, 12.4%) |  | 727 | 166 | 22.8% (19.9%, 26.0%) |  |
| <= 3 days (High) | 180 | 11 | 6.1% (3.4%, 10.6%) |  | 180 | 30 | 16.7% (11.9%, 22.8%) |  |
| 4+ days (Low) | 371 | 52 | 14.0% (10.9%, 17.9%) |  | 371 | 118 | 31.8% (27.3%, 36.7%) |  |
| 4+ days (Medium) | 1,279 | 160 | 12.5% (10.8%, 14.4%) |  | 1,279 | 383 | 29.9% (27.5%, 32.5%) |  |
| 4+ days (High) | 335 | 35 | 10.4% (7.6%, 14.2%) |  | 335 | 85 | 25.4% (21.0%, 30.3%) |  |

